# Protective Effects of Increased EHR-Mediated Information Sharing Within- and Between-Healthcare Teams on Patient Outcomes

**DOI:** 10.64898/2026.09.22.26362777

**Authors:** Shin-Ping Tu, Daniel Sewell, Brittany Garcia, Quinn Stoddard O’Neill, Michael Hogarth, Marissa Shuffler, Aaron Boussina, Helen Chew, David Cooke, Patrick Romano, Joann Elmore, Alan Dow, Xi Zhu

**Affiliations:** University of California Davis, Sacramento, California, U.S.A; University of Iowa, Iowa City, Iowa, U.S.A; University of San Diego, La Jolla, California, U.S.A; Clemson University, Clemson, South Carolina, U.S.A; University of California Los Angeles, Los Angeles, California, U.S.A; Virginia Commonwealth University, Richmond, Virginia, U.S.A

## Abstract

Electronic Health Records (EHRs) provide a common repository for patient information, and while many challenges persist, EHRs allow asynchronous coordination among healthcare professionals (HCPs). Such coordination is inherently challenging, as large multidisciplinary teams, or teams-of-teams, of HCPs jointly manage complex patient care. These teams-of-teams naturally fit under the Multiteam System (MTS) framework, where evidence suggests that both stronger within- and between-team communication improves overall team performance. Yet little is known how about these patterns of information sharing mediated through the EHR correlate with patient outcomes. We extracted over 53.9 million EHR access logs for 482 patients to construct novel within- and between-team information sharing measures which were then used to predict future emergency department (ED) visits and hospitalizations admitted through the ED. These network measures showed both statistical and practical significance, suggesting that this modifiable dimension of teamwork-information sharing through the EHR-could reduce acute care utilization.

## Introduction

Electronic Health Records (EHRs) are now ubiquitous in the US,^1^ and increasingly serve as a platform for digital communication and coordination among healthcare professionals as well as with patients and caregivers through patient portals.^2–6^ As information repositories, EHRs: (i) provide the ability of information holders and information retrievers to carry out their tasks asynchronously; (ii) reduces information processing load in a team because information holders may satisfy multiple requests from information retrievers through a single contribution to the repository; and (iii) enable direct access to “external” information from outside the team when multiple teams are all active users of the common repository.^7, 8^

However, computer-mediated communication by distributed teams also present significant challenges to information sharing for complex and interdependent tasks.^9–11^ The body of literature documenting the role of EHRs in patient care inefficiencies, errors, and physician burnout continues to grow,^12–16^ with a report from the U.S. Department of Health and Human Services (HHS) highlighting major issues and concerns related to EHR burden including clinical documentation as well as the usability and user experience of health information technology (IT).^17^ As research supporting these pain points expand, HHS provided guidance for actions to implement and address these issues: reducing the regulatory burden of documentation; developing a process to address inconsistent data collection by federal, state, and local programs standardizing service orders and data; improving the user interface to match clinical workflow; and improving the value and usability of electronic clinical quality measures.^17^

As the volume and complexity of EHR data continue to grow rapidly,^18, 19^ our team embarked to study the complex information sharing networks of healthcare professionals (HCPs) engaged in patient care through the EHR. These networks are comprised of HCPs in multiple specialties needing to communicate important patient information both with others in the same specialty team as well as with those in different specialty teams. Such complex systems naturally fit into the Multiteam System (MTS) framework from the organizational psychology field.^20, 21^

To achieve better performance, MTS theory submits that the system must develop effective communication and coordination within and between teams.^22^ These guiding MTS principles have been found salient in healthcare teams delivering complex care to patients. Increased HCP communication has been shown to improve patient outcomes and overall team decision making,^23^ and conversely poor communication has been shown to be a leading cause of medical errors^24^. Notably researchers have found significant associations between patient outcomes and both within and between specialty teamwork and communication.^25, 26^

Our research centers on one modifiable dimension of teamwork: information sharing through EHRs. MTSs, or teams-of-teams, are networks of interdependent teams, with collective system goals (e.g., patient-centered care and overall quality) in addition to local team goals (e.g., discipline-specific quality measures).^27–30^ While interpersonal communication amongst MTS members is restricted by physical design and proximity, EHRs transcend physical constraints to allow for within-team and between-team information sharing. Thus, while significant challenges remain endemic in EHR systems, the potential still exists for large teams-of-teams to asynchronously engage HCPs within and between teams in complex patient care for improved outcomes.

Cancer is a complex disease requiring multiple specialties to coordinate care, thereby creating potential for varied quality in within and between team communication. Cancer patients are also highly vulnerable to adverse health outcomes,^31^ thus an important population to study the relationship between EHR-mediated information sharing and patient outcomes. Our research focuses on cancer patients and EHR-mediated information sharing amongst HCPs participating in their care, and this paper reports the results from our study testing these two hypotheses:

**Hypothesis 1**. Increased information sharing within teams is associated with decreased risk for emergency department (ED) visits and hospitalizations.

**Hypothesis 2**. Increased information sharing between teams is associated with decreased risk for ED visits and hospitalizations.

## Methods

### Study Population

The University of California Institutional Review Board approved our research (IRB #1988738) with patients 18 years and older diagnosed with stage II and III breast, colorectal, or non-small cell lung cancers from January 1, 2016, to December 31, 2021. We identified eligible patients from the University of California, San Diego (UCSD) cancer center registry and restricted our study sample to patients with stage II and III cancer based on the intensive and multidisciplinary coordination required for the treatment of these cancer stages.^32, 33^ Patients with the same diagnosis dates that included an ineligible cancer stage were excluded (e.g. breast cancer stage I and II both diagnosed on July 3, 2017). Moreover, we only included patients who were diagnosed and received their initial course of treatment at UCSD (class of case 12 and 14).

### Study Perio

The index date for each study patient was determined by the first time they were diagnosed with stage II or III cancer. Our study period started four weeks prior to the index date and continued until 12 weeks after the index date. To be included in this study, patients had EHR access log data for the period ranging between four weeks prior through 8 weeks past the index date (see *Electronic Access Logs* below) and outcomes data ranging between 9 to 12 weeks after the index date (see *HCP Data* below). We excluded patients who died, had a second cancer site diagnosis before 12 weeks post index date, or had a date of last contact with the UCSD health system before 12 weeks post index date.

### Patient Data

EHRs from two hospitals of the UCSD health system provided these patient data: age at diagnosis, sex (male or female), racialized group, Spanish origin, marital status at diagnosis, insurance payer, and 34 non-cancer comorbidities identified using the Elixhauser methodology.^34^

### HCP Data

We extracted EHR access log data of each patient from one year prior to two years after their first cancer diagnosis. Based on information of users accessing the EHR and their specific actions (e.g., modify or view clinical notes) we identified: (1) which HCPs were participating in a patient’s care and when; as well as (2) information sharing between HCPs through authoring and viewing clinical notes.

For each patient, we captured each HCP who accessed that patient’s records, recording the first and last access log associated with that patient and that HCP; this gave us the period during which that HCP was engaged in that patient’s care. For each week from the index date through the eighth week, we recorded which HCPs were active, and thus which MTS component teams (see *MTS Component Team Assignations* below) were active.

We also filtered the access logs to only those actions related to clinical notes, excluding billing-related notes and notes which were eventually deleted. When a scribe was the note author, we took the next HCP to modify the note and attributed the authorship to that HCP at the scribe’s timestamp. Similarly, when a medical student modified a note, we attributed authorship to the physician who signed off on the note. When a note had multiple modifiers, we considered subsequent modifying HCPs to have viewed the information written by past modifiers, hence leading to information both being shared from past modifiers to the HCP and from the HCP to future viewers (and future modifiers).

### MTS Component Team Assignations

EHR access logs include data on provider specialty (e.g., Anesthesiology), provider type (e.g., Neurophysiology Tech), and clinician title (e.g., LVN/LPN). We assigned HCPs to twenty derived MTS component teams by provider specialty first. If a HCP was missing a specialty, their provider type was used. When both provider specialty and provider type were missing, then clinician title was used.

HCPs who did not have any information on provider specialty, provider type nor clinician title were excluded from the analyses. We also excluded non-clinical professionals (i.e., clinical care affiliates and partners, coders, data coordinators, financial counselors, and front desk personnel), scribes, and students.

### Outcome Data of ED Visits and Hospitalizations

The California Department of Health Care Access and Information (HCAI) provided patient ED visits and hospitalizations from January 1, 2015, through Dec 31, 2022. For this study, we only included hospitalizations when patients were admitted through EDs. Multiple ED visits or hospitalizations on the same day to the same facility were considered a single visit/hospitalization. For each patient we had two outcomes of interest: the number of ED visits during weeks 9 to 12 post index date, and the number of hospitalizations during weeks 9 to 12 post index date.

### Network Construction

For each patient, we constructed a directed, bipartite, dynamic network. Each edge connected either a HCP to a note if the HCP engaged in an authorship activity, or else a note to a HCP if the HCP viewed a note or modified a note that had an earlier, different note modifier. Figure 1 gives an example of a particular patient’s first four weeks after their index date (isolates, that is, unconnected notes or HCPs, are dropped for visualization purposes). While not depicted in Figure 1, it is important to bear in mind that the HCPs in the network are continually changing as they become active or cease to take part in a patient’s care.

**Figure 1.**
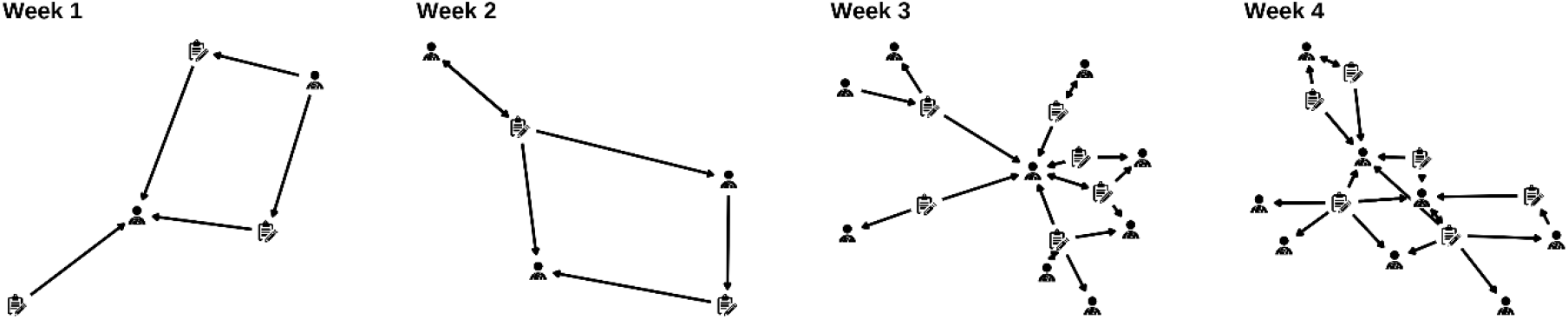
An example patient’s bipartite network during the first four weeks post their index date. Nodes of the network are either HCPs or notes and are represented by their respective icons. HCPs connect to nodes and vice versa, but nodes of similar type do not connect directly to each other.

From a patient’s bipartite network and HCP assignations to MTS component teams, we constructed the following two network measures for each week post index date up to 8 weeks. First, for each MTS component team, we computed the <u>within-team information sharing</u> measure, defined as the number of notes written by a member of the MTS component team and read by different member of the same MTS component team. If no team members were active or if no other team members besides the author was active during the given week for the given patient, we set the within-team information sharing value to be not applicable (NA). Similarly, for each ordered pair of MTS teams, we computed the <u>between-team information sharing</u> measure, defined to be the number of notes written by a member of the first MTS team of the pair (source team) and read by a member of the second MTS team of the pair (destination team). As before, if no member of the destination team was active during the given week for the given patient, we set the between-team information share for this ordered pair to be NA.

To create a single patient-level network measure for each team (for within-team share) or ordered pair of teams (for between-team share), we took the average of each non-NA valued network measure. In addition, we similarly recorded the average team size for each patient, where the team sizes were averaged only over those weeks where at least one HCP from that team was actively participating in the patient’s care. If a patient never had a MTS team represented (e.g., if a patient never received mental health care), the patient’s average team size for that team was stored as NA.

### Statistical Analysis

To investigate the association between the within-team and between-team information sharing measures and the number of ED visits and hospitalizations, we fit a sequence of generalized linear models (GLMs) based on the Poisson distribution. Again, the network measures were constructed over the time interval of 0 to 8 weeks post index date, and the outcome healthcare encounters were counted over the period from 9 to 12 weeks post index date. In our GLMs, we controlled for cancer site, cancer stage, age, and each of the 34 comorbidities that contribute to the Elixhauser comorbidity index. We additionally controlled for the MTS team sizes in the GLMs to ensure that our network measures were not simply acting as a proxy for the presence or overall activity of a given MTS team.

We fit a separate model for each network measure and each outcome due to two factors. First, the number of patients which had non-NA values for a given network measure varied greatly across teams and ordered pairs of teams. Second, the total number of potential network measures that could be considered was 400 (=20 within-team + 20×19 between-team measures) and combined with the number of confounding covariates would produce nearly as many covariates as observations, leading to high numerical instability, statistical non-estimability, and extremely low power.

Due to the low counts of our outcomes and network measures with high counts of patients with NA values, several of our network measures were non-estimable with our sample size. We only fit GLMs where when we created a 2×2 contingency table consisting of non-zero outcomes (0, > 0), and non-zero network measure (0, > 0), the minimum cell count was 5 or greater. Additionally, for each GLM we only included the comorbidities which when similarly tabulated also had a cell count of 5 or more.

Inference was made within the Bayesian statistical framework. Statistical significance was determined through the use of Bayes factors, using the guidance of Kass and Raftery ^35^ to determine substantial evidence in favor of the inclusion of the network measure in the GLM. We also computed point estimates via the posterior mean, 95% credible intervals, probability of direction, and the posterior probability that the effect lay in the region of practical equivalence (ROPE). The probability of direction is a key quantity in Bayesian inference which states the degree of certainty that the direction of the effect is known. In our context where the rates of ED visits or hospitalizations are being modeled, rate ratios less than 1 indicate a protective effect due to an increase in the covariate, and rate ratios greater than 1 indicate a harmful effect. Thus, the probability of direction is the degree to which we are certain an effect is either protective or harmful.

The ROPE, which captures *practical* significance rather than just *statistical* significance, was determined in a fashion similar in logic to the guidance provided by Kruschke ^36^ If a shift across the full range of a network measure led to less than half the size of a small effect in the rate of the outcome, we considered this to be a practically meaningless effect size. More specifically, considering a small effect size to be a rate ratio of 1.25 (or its reciprocal), this led to a ROPE which bounded the GLM regression coefficient in absolute value by log(1.125) /4*ss*_*xx*_, where *ss*_*xx*_ is the standard deviation of the network measure covariate. Smaller values of the posterior probability that the true effect is in the ROPE indicates that we are more certain that the true effect is practically meaningful. All analyses were done in R (v4.5.2)^37^ using the *bayesics* R package.^38^

## Results

We identified 7,990 patients with breast, colorectal, and non-small cell lung cancer at UCSD Health from January 1, 2016 to December 31, 2021. After applying the inclusion criteria, our study sample consisted of 482 cancer patients. Table 1 provides the demographic characteristics of our study sample, disaggregated by cancer site. Figure 2 shows the percent of patients with each comorbidity in our analyses. We extracted over 53.9 million EHR access actions performed on our study patients with 88,134 unique notes written.

**Table 1.**
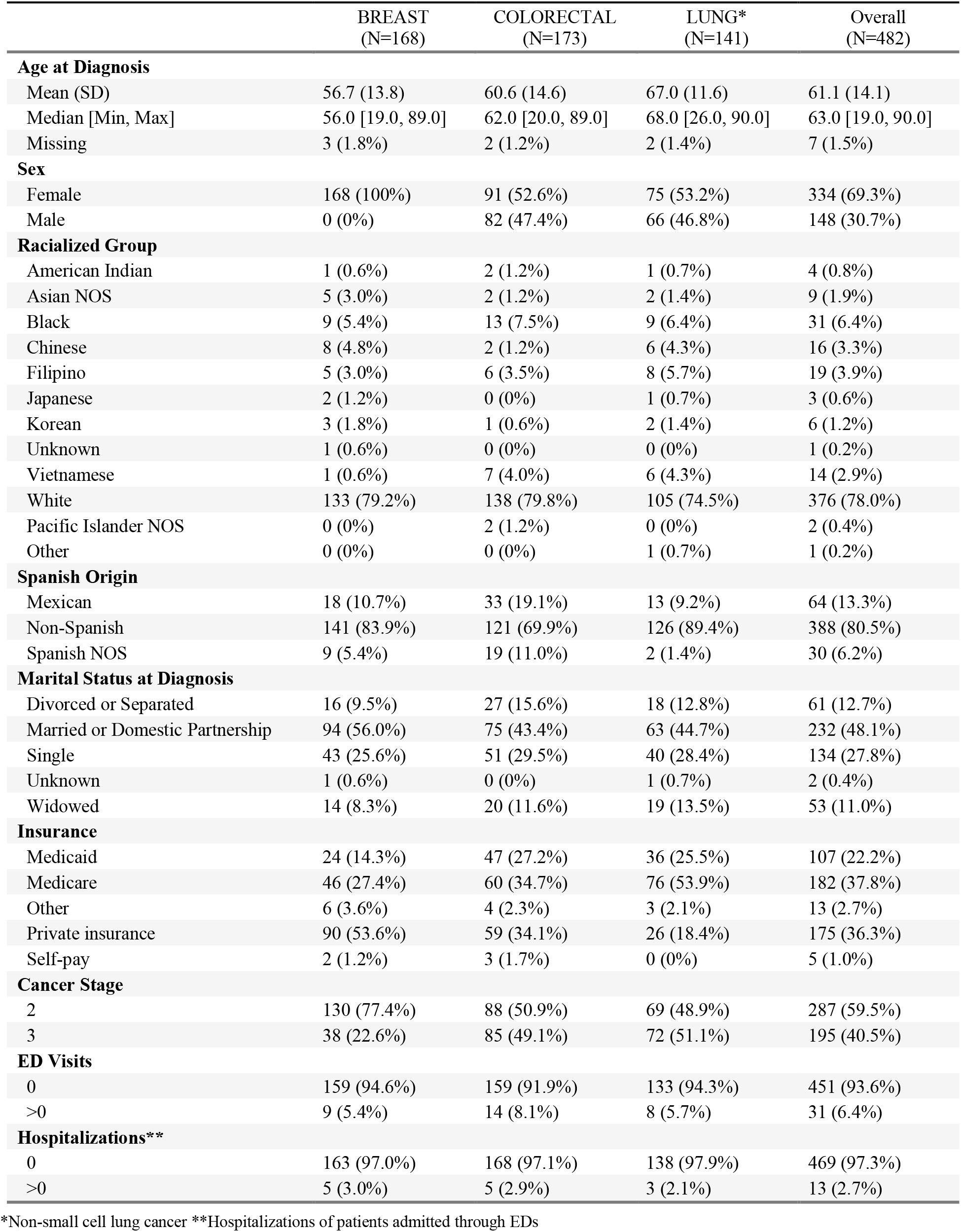
Cancer patient characteristics.

**Figure 2.**
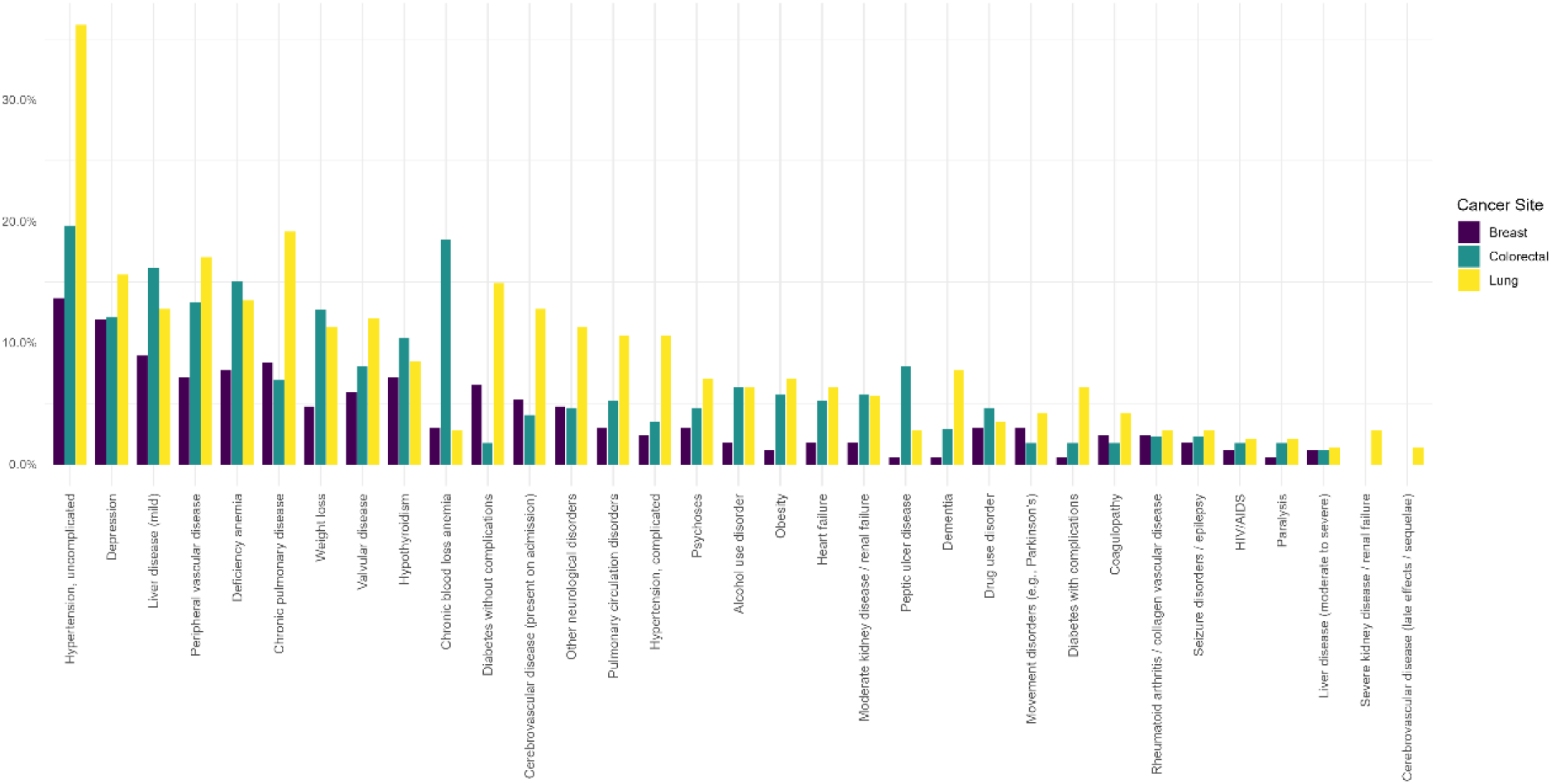
Prevalence of comorbidities in study patients by cancer site.

Table 2 provides the average and standard deviation of team size for all 20 MTS component teams. As described above, these summary statistics are taken over only those weeks in which at least one member from that MTS team was active in that patient’s care. Unsurprisingly, the nursing team was the largest, followed by the pharmacy team with 19.8 team members on average. General practice, pediatrics, radiation oncology, and pathology had the smallest teams, with an average size near 1.

**Table 2.** Association of within-in team information sharing with patient outcomes of ED visits and hospitalizations admitted through the ED. Statistically significant results, as determined by Bayes factors displaying substantial evidence in favor of an association, are bolded. Non-estimability is indicated by ---.

| MTS Component Teams |  | Patient Outcomes |  |  |  |  |  |
| --- | --- | --- | --- | --- | --- | --- | --- |
| Teams | Size<br>(#HCPs) | ED Visits |  |  | Hospitalizations |  |  |
|  | Mean (SD) | Estimate<br>(95% CI) | Prob. Dir. | Pr(ROPE) | Estimate<br>(95% CI) | Prob. Dir. | Pr(ROPE) |
| Ancillary | 6.48 (7.25) | --- | --- | --- | --- | --- | --- |
| Dietary | 1.89 (1.56) | 0.797<br>(0.549,1.158) | 0.88 | 0.04 | --- | --- | --- |
| Emergency Medicine | 3.43 (4.4) | 1.089<br>(0.735,1.614) | 0.67 | 0.07 | --- | --- | --- |
| Family Practice | 1.74 (1.27) | --- | --- | --- | --- | --- | --- |
| General Practice | 1 (0) | --- | --- | --- | --- | --- | --- |
| Internal Medicine | 2.57 (3.23) | 0.933<br>(0.726,1.198) | 0.71 | 0.09 | --- | --- | --- |
| Internal Medicine Subspecialty | 2.63 (3.21) | **0.854**<br>**(0.757,0.964)** | **0.99** | **<0.001** | --- | --- | --- |
| Medical Oncology | 1.68 (1.05) | 1.136<br>(0.828,1.558) | 0.79 | 0.10 | --- | --- | --- |
| Mental Health | 1.53 (0.85) | --- | --- | --- | --- | --- | --- |
| Nursing | 26.58 (25.59) | --- | --- | --- | --- | --- | --- |
| Pathology | 1.3 (0.65) | --- | --- | --- | --- | --- | --- |
| Patient Support Services | 3.97 (4.32) | 1.045<br>(0.996,1.096) | 0.96 | 0.05 | --- | --- | --- |
| Pediatrics | 1.01 (0.12) | --- | --- | --- | --- | --- | --- |
| Pharmacy | 10.78 (10.94) | **0.753**<br>**(0.623,0.911)** | **>0.99** | **<0.001** | --- | --- | --- |
| Radiation Oncology | 1.18 (0.46) | --- | --- | --- | --- | --- | --- |
| Radiology | 4.64 (3.77) | --- | --- | --- | --- | --- | --- |
| Specialty Other | 2.92 (3.32) | 1.011<br>(0.991,1.032) | 0.86 | 0.14 | **0.917**<br>**(0.9,0.935)** | **>0.99** | **<0.001** |
| Surgery Other | 1.63 (1.17) | --- | --- | --- | --- | --- | --- |
| Surgical Oncology | 2.84 (2.95) | 0.986<br>(0.891,1.091) | 0.61 | 0.12 | --- | --- | --- |
| Therapy | 5.23 (9) | 0.663<br>(0.439,1.003) | 0.97 | 0.01 | --- | --- | --- |

Results from our analyses examining the relationship of within-team information sharing and our two outcomes of interest, ED visits and hospitalizations are also listed in Table 2. Ten MTS teams had estimable within-team network measures for ED visits. Of these, Internal Medicine Subspecialty and Pharmacy teams had statistically significant associations with ED visit rates. Both these effects were protective, implying that higher levels of information sharing within these teams led to a decrease in ED visit rate. When modeling the associations of within-team network measures and hospitalizations, only the Specialty Other MTS team was estimable; in large part, this was due to the sparsity of this outcome (see Table 1). This effect was also statistically significant and indicative of a protective effect from increased information sharing amongst team members. Further, all three of these effects had negligible posterior probabilities that the true effects were in the region of practical equivalence (ROPE), implying that these statistically significant results were also practically meaningful with very high probability.

There were 72 between-team information sharing measures that were estimable when modeling ED visit rates and 9 when estimating hospitalization rates. Of these, four were statistically significant, all of which corresponded to ED visits. For brevity, we have only included these four results in Table 3. All four effects were protective, indicating that a higher level of information sharing from the authoring team to the viewing team reduced the rate of ED visits. As with the within-team measures, all four effects have very low posterior probability of being in the ROPE, indicating that these statistically significant results are also practically meaningful with high probability.

**Table 3.** Association of between-team information sharing with patient outcomes of ED visits. All results shown are statistically significant as determined by Bayes factors displaying substantial evidence in favor of an association, are bolded.

| MTS Component Teams |  | Patient Outcomes ED Visits |  |  |
| --- | --- | --- | --- | --- |
| Author | Viewer | Estimate (95% CI) | Prob. Dir. | Pr(ROPE) |
| Internal Medicine Subspecialty | Nursing | **0.817 (0.682,0.979)** | 0.986 | 0.005 |
| Internal Medicine | Specialty Other | **0.803 (0.684,0.943)** | 0.996 | 0.002 |
| Nursing | Internal Medicine Subspecialty | **0.72 (0.535,0.969)** | 0.985 | 0.005 |
| Specialty Other | Internal Medicine Subspecialty | **0.671 (0.49,0.918)** | 0.994 | 0.003 |

## Discussion

To our knowledge, this study is among the first to examine clinical MTSs using real-world healthcare delivery data derived from EHR access logs and to operationalize MTS theory through quantitative network measures of EHR-mediated information sharing. While prior literature has largely focused on documentation burden, usability challenges, and clinician burnout, our work shifts attention toward how HCPs access and share clinical information in the EHR to coordinate care for patients with complex health conditions.

Our findings demonstrate that greater information sharing within specific teams was associated with improved patient outcomes, reflected in lower rates of ED visits and hospitalizations for certain teams. Similarly, increased between-team information sharing was associated with fewer ED visits. These results are consistent with MTS-informed care coordination research emphasizing cross-boundary integration as a determinant of healthcare performance.^28^

Importantly, our results are consistent with the Multitheoretical Multilevel communication model^39^ that effective information sharing is structural and not simply based on quantity or frequency of information sharing. Prior patient safety research cautions that excessive or poorly organized information can contribute to cognitive overload and diagnostic risk.^16^ The protective associations observed in our study support meaningful information exchange and coordination rather than indiscriminate increases in EHR activity, and underscore the value of network-based conceptual frameworks and metrics to capture the relational structure of complex patient care.

This study adds to the quality and safety literature and foundational work to advance learning health systems using EHR audit log data.^40–43^ National leaders in clinical informatics have called for the transformation of routinely collected digital data into scalable indicators of care delivery performance. Our research aligns with the National Committee for Quality Assurance (NCQA) call for a change to fully digital quality measure frameworks by 2030, with a shift away from administrative and claims data towards leveraging EHR data to provide a more real-world reflection of quality of care.^44^ More broadly, our work supports learning health systems strategies that emphasize leveraging routinely generated digital health data to enable continuous evaluation, rapid-cycle improvement, and scalable innovation in care delivery.

### Limitations

Several limitations warrant consideration. First, this was a single-site study conducted within an academic health system. Institutional culture, team configuration, and EHR customization may influence both information sharing patterns and outcomes. Second, we aggregated three cancer sites in the analyses. Although all three cancers involve multidisciplinary coordination, we acknowledge that disease-specific workflows differ and plan to stratify by cancer site as part of our analysis with data from three sites (UC Davis, UC Los Angeles and UCSD). Third, although we included patients diagnosed and treated at UCSD Health, this analysis did not adjust for patients who received primary care at UCSD Health. Fourth, in our early conceptualization of MTS component teams using EHR access log data, we aggregated HCPs into MTS component teams that may not adequately represent the clinical group they belong to (i.e., Internal Medicine Subspecialty; Nursing, Specialty Other teams). We also chose to keep Family Medicine, General Practice, and Internal Medicine as separate MTS component teams instead of combining them into a Primary Care component team. Fifth, although this study did not distinguish inpatient from outpatient settings, we plan to do so for future analyses. Finally, while access-log data objectively capture interaction events, they do not directly assess content quality, accuracy, or interpretive alignment among team members. As prior work on diagnostic safety and information overload suggests, the structure of communication is only one component of safe information exchange.^16^

### Future directions

Future work can extend this research in several ways. First, multi-site analyses with two additional academic health systems will determine the generalizability of the findings from this single site study. Second, stratification by cancer site and stage will clarify disease-specific information sharing and coordination patterns. Third, distinguishing inpatient and outpatient teams will allow detailed examination of transitions of care. Fourth, incorporating note type-specific analyses may improve specificity by focusing on high-value clinical documentation, as well as identifying network structures vulnerable to break downs in the flow of information. Fifth, refinement of HCP assignations (e.g., Internal Medicine Subspecialty team) to more specific component teams (e.g., Cardiology, Gastroenterology, Nephrology) may provide additional insights of between team information sharing and patient outcomes. Fifth, extending our analysis to unplanned hospitalizations will align this research with the cancer literature. Sixth, the network measures developed from our research will be applicable as healthcare delivery increasingly evolves towards Human-AI teams. Most importantly, we envision this line of research will contribute significantly to the advancement of EHR data visualization of that can reduce cognitive load, support working memory, and potentially reduce physician workload while enhancing patient care.

### Conclusion

Our use of EHR access logs aligns with prior validation work demonstrating that audit log data can reliably characterize clinical work patterns and workflow behaviors.^45, 46^ Whereas much of the existing audit log literature has focused on measuring documentation time or clinician workload, our research extends the application of conceptual frameworks and methodologies for Learning Health Systems in the Human-AI age.

## Data Availability

The patient-level data used in this study are not publicly available due to institutional, privacy, and data-use restrictions. UCSD electronic health record, cancer center, and EHR access-log data cannot be publicly shared. Data from the California Department of Health Care Access and Information (HCAI) and California Cancer Registry (CCR) were obtained under data-use agreements that prohibit redistribution outside the authorized research team. Therefore, the analytic datasets cannot be deposited in a public repository or shared directly by the authors. Access to these data requires appropriate approvals and agreements with the respective data custodians.

## Acknowledgement

Our research is supported by grant R01CA273058 from the National Cancer Institute. Contents of this manuscript are solely the responsibility of the authors and do not represent the official view of the National Cancer Institute.

